# A Machine Learning Approach to Differentiate Between COVID-19 and Influenza Infection Using Synthetic Infection and Immune Response Data

**DOI:** 10.1101/2022.01.27.22269978

**Authors:** Suzan Farhang-Sardroodi, Mohammad Sajjad Ghaemi, Morgan Craig, Hsu Kiang Ooi, Jane M Heffernan

## Abstract

Data analysis is widely used to generate new insights into human disease mechanisms and provide better treatment methods. In this work, we used the mechanistic models of viral infection to generate synthetic data of influenza and COVID-19 patients. We then developed and validated a supervised machine learning model that can distinguish between the two infections. Influenza and COVID-19 are contagious respiratory illnesses that are caused by different pathogenic viruses but appeared with similar initial presentations. While having the same primary signs COVID-19 can produce more severe symptoms, illnesses, and higher mortality. The predictive model performance was externally evaluated by the ROC AUC metric (area under the receiver operating characteristic curve) on 100 virtual patients from each cohort and was able to achieve at least AUC=91% using our multiclass classifier. The current investigation highlighted the ability of machine learning models to accurately identify two different diseases based on major components of viral infection and immune response. The model predicted a dominant role for viral load and productively infected cells through the feature selection process.

## 1 Introduction

Severe acute respiratory syndrome coronavirus 2 (SARS-CoV-2) and influenza viruses cause COVID-19 and influenza diseases, respectively, and mainly infect the upper and lower respiratory tract [1, 2]. Both infections present some similar prime symptoms leading to a clinical dilemma in diagnosing patients with the early infections [3–5]. However, COVID-19 tends to cause worse decompensation due to its intensive transmission, and vascular effects which have led to an unrivaled global crisis [6–9]. Moreover, as the striking COVID-19 outbreak continues, the concurrence of epidemics can be impendent. Therefore, it is of interest to design data analysis tools that can accurately differentiate between these two infections and help curb the pandemics.

One way to rapidly classify patients as influenza or COVID-19 could be through machine learning approaches. Preliminary investigation illustrated the potentials of machine-learning models for accurately distinguishing between these two viral infections, using demographics, body mass index, and vital signs in infected patients [9]. Herein, we used a simple ML-based classification to identify the patients with influenza and SARS-CoV-2 using mathematically based variables of the in-host infection dynamics and immune response. During the past decade, virus-host mathematical modeling has become an increasingly powerful tool to study intracellular viral infection dynamics and the ensuing immune response. Dynamic mathematical modeling can deepen our understanding of virus spread within organs that amplify the development of new antiviral drugs, and optimize treatment regimens. Importantly, these models can also help to mitigate difficulties related to clinical data analyses, such as inconsistencies in data collection that can lead to biased trial results and significantly complicate comparative analytics. For this purpose, we applied a basic mathematical model on the cellular scale (the so-called target cell-limited model [10, 11]) fit two different sets of *in vivo* data, to create virtual patient cohorts. Using our provided multiclass classifier, the patients were differentiated between the two infections. We certainly hope that our work can be a guide for future applications validated on the external clinical test set to help clinicians and front-line healthcare workers accurately recognize the disease. With just some important in-host measurements clinicians may discern the patients before a laboratory diagnosis.

This paper is organized as follows: In section 2, through subsection 2.1, we discuss the In-host mathematical modelling of influenza and COVID-19 and parameter estimation. Via subsection 2.2, we use the mechanistic model to generate synthetic patient data. In subsections 2.3 we study developing and evaluating a supervised machine learning method to discriminate the patients with different infections. The Interpretability of the developed model is discussed in subsection 2.4. The results of the prediction are presented in section 3 through subsection 3.1. Subsection 3.2 discusses the importance of the data features and determines the dominant features. The paper concludes with a discussion in Section 4.

## 2 Method

### 2.1 Mechanistic models

We employed a target-cell limited model of viral dynamics using five differential equations that track susceptible target cells (*T*), infected cells in the eclipse phase (*I*_1_), productively infected cells (*I*_2_), virus (*V*), and interferon (*F*) in-host. Figure 1 presents a flow diagram of the model. The system of ordinary differential equations is as follows:

**Figure 1:**
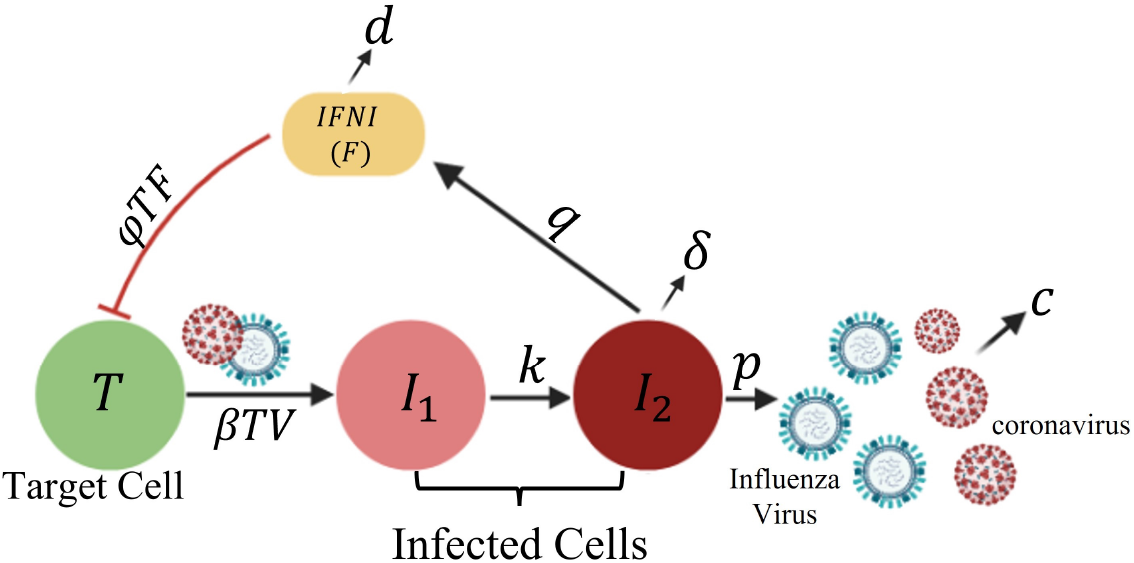
Schematic of viral infection. Each Target cell, T, is infected by a virus, V, with a constant rate *β*. During the eclipse period the productively infected cell, *I*_2_, is being produced by the first infected cell, *I*_1_, with a constant rate *k*. The Infected cell, *I*_2_, produces virus at rate p, IFNI at rate q and dies at rate *δ* per cell. IFNI hinders viral infection by converting target cells to a virus-resistant state with a constant rate *ϕ* and decays with rate *d*. Free virus particles that can be influenza or coronaviruses are cleared at per-capita rate c.

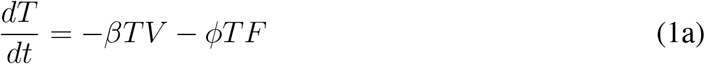

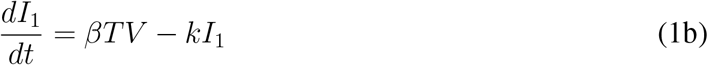

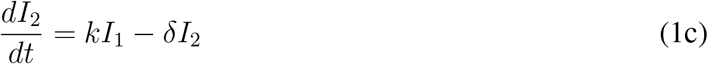

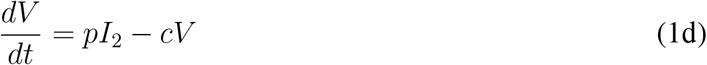

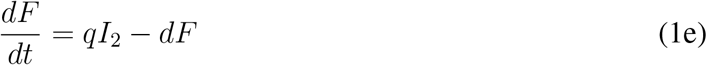

Briefly, virus particles *V* can infect susceptible target cells *T* to produce infected cells. This is represented by the term *βTV*. Newly infected cells first enter the eclipse phase *I*_1_ and become productively infected cells *I*_2_ when within-cell processes that program the cell to make new virus particles are completed. The eclipse phase takes, on average, 1*/k* time units. Productively infected cells produce new virus particles with a rate of *p*, and the virus particles are cleared from the system with a rate of *c*. We assumed that productively infected target cells have a death rate *δ*. Susceptible target cells can be protected from infection by Type I interferon (IFNI), *F*. Type I interferons protect neighboring cells from infection and elicit an immune response [12, 13]. They are central to combating different virus infections and are regularly measured in clinical trials or infection studies in humans and animals [14]. We assumed that interferon production is proportional to the number of productively infected cells, that interferon has a natural decay rate *d*, and that interferon protects susceptible cells by removing them from the susceptible target cells population, with a rate *ϕF*. This term was ignored in [10] for influenza infection. The model described by Eq. 1 was used in [10] and [11] to examine the kinetics of influenza A and SARS-CoV-2 viral dynamics, respectively. For the sake of simplicity, we have ignored a half-day lag in IFNI response that was considered in [10].

#### 2.1.1 Parameter Estimation

Model parameters for influenza A infection were fit to data from an experimental H1N1 influenza A/Hong Kong/123/77 infection for six patients [10] and for SARS-CoV-2 from thirteen untreated patients infected with severe acute respiratory syndrome-coronavirus [11]. The geometric average parameter values along with their 95% confidence intervals and units are summarized in Table 1. We assumed that the initial number of target cells, *T*_0_, is equal to the total number of target cells in the upper respiratory tract and set *T*_0_ = 4 *×* 10^8^ cells. In [11] the authors considered that the target cells distributed in a volume of 30 mL. Assuming that 1% of these cells expresses the angiotensin-converting enzyme 2 (ACE2) as a receptor for SARS-CoV-2, the target cell concentration, *T*_0_, was expressed as 1.33 *×* 10^5^ cell/ml. Model variables with initial values were estimated as in Table 2.

**Table 1:** Average values and confidence intervals, *CI*, for influenza A and SARS-CoV-2 withinhost viral infection model parameters. Confidence levels of 95% displays the degree of certainty that the parameter values for different samples, fall around the mean.

| Influenza Model Parameters [10] |  |  |  |  |  |  |  |  |  |
| --- | --- | --- | --- | --- | --- | --- | --- | --- | --- |
| $V_0$ [95%CI]<br>$TCID_{50}/ml^1$ | $R_0$ | $\beta$ [95%CI]<br>$(TCID_{50}/ml)^{-1}d^{-1}$ | $k$ [95%CI]<br>$d^{-1}$ | $p$ [95%CI]<br>$(TCID_{50}/ml)d^{-1}$ | $c$ [95%CI]<br>$d^{-1}$ | $\delta$ [95%CI]<br>$d^{-1}$ | $q$<br>$d^{-1}$ | $d$<br>$d^{-1}$ | $\phi$<br>$d^{-1}cell^{-1}$ |
| 0.075[7.6E - 4, 7.5]<br>SD:3.5724 | 21.5[10.1-46.1]<br>SD:17.15 | 3.2E - 5[6E - 6, 1.7E - 4]<br>SD:7.8124 | 4[3, 5.2]<br>SD:1.0486 | 0.046[0.012, 0.17]<br>SD:0.07527 | 5.2[3.1 - 8.7]<br>SD:2.6677 | 5.2[3.2 - 8.6]<br>SD:2.5724 | 1 | 1.9 [12, 15, 16] | 0 |
| COVID-19 Model Parameters [11] |  |  |  |  |  |  |  |  |  |
| $V_0$<br>$Copies/ml$ | $R_0$ 95%[CI] | $\beta$<br>$(Copies/ml)^{-1}d^{-1}$ | $k$<br>$d^{-1}$ | $p$ 95%[CI]<br>$(Copies/ml)d^{-1}$ | $c$<br>$d^{-1}$ | $\delta$ 95%[CI]<br>$d^{-1}$ | $q$<br>$d^{-1}$ | $d$<br>$d^{-1}$ | $\phi$<br>$d^{-1}cell^{-1}$ |
| 0.1 | 8.6[1.9 - 17.6]<br>SD:12.9893 | 5.68E - 9 | 3 | 22.71[0 - 59.64]<br>SD:49.3426 | 10 | 0.6[0.22 - 0.97]<br>SD:0.62051 | 1 | 0.4 | 1.97E-6 [17] |

<sup>1</sup> 1 $[TCID_{50}/ml]$ corresponds to 4000 $[Copies/ml]$ [18].
<sup>2</sup> $R_0$ is the basic reproduction number.

**Table 2:** Model Variables with Initial values.

| Variable | Definition | Initial Value | Unit |
| --- | --- | --- | --- |
| $T$ | Target cell | 4E+8 | Cell |
| $I_1$ | Infected cell (eclipse phase) | 0 | Cell |
| $I_2$ | Productively infected cell | 0 | Cell |
| $V$ | Viral load (flu) | 7.5E-2 | $TCID_{50}/ml$ |
| | (COVID-19) | 0.1 | $Copies/ml$ |
| $F$ | type I interferon (IFNI) | 0 | Interferon |

### 2.2 Generation of Virtual Patients

To generate a cohort of 100 virtual patients, we followed a technique similar to the one used in [13]. Initial parameter sets representing individual virtual patients were drawn from normal distributions with means fixed to the corresponding parameter value in Table 1 and standard deviations derived from confidence interval measurements. Standard deviations were obtained from standard errors, confidence intervals, and *t* statistics which measure the size of the difference relative to the variation in the sample data. For each parameter value, the standard deviation was obtained by dividing the length of the confidence interval by standard errors width (2 ×*t* − *value*) and then multiplying by the square root of the sample size as follows

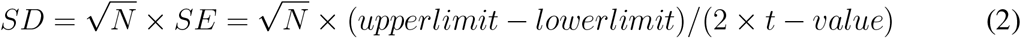

Standard errors must be of means calculated from within each parameter confidence interval. The *t* − *value* for a 95% confidence interval from a sample size of *N* was then obtained in Microsoft Excel using the *tinv* function (i.e. *tinv*(1 − 0.95, *N* − 1)). From [10], the sample size for the influenza cohort is 6 patients infected by H1N1 influenza A/Hong Kong/123/77 infection. The COVID-19 cohort consisted of 13 untreated patients infected with severe acute Respiratory syndrome-coronavirus2 [11]. Therefore, the *t* − *value* for influenza patients is 2.571 and for COVID-10 patients is about 2.179. From normal distributions with standard deviations, *σ*, and means, *µ*, as the original parameter values, we then generated normal distributions covering values lying around each parameter value such that | *µ* ± *σ* − *µ*| *< h*. Herein, the parameter *h* is the user-defined value as a measure of data diversity. In the other words, the bigger the parameter *h*, the more diverse the synthetic data. Accordingly, the external noise can affect the data through the parameter *h*. The dynamics of 100 virtual patients from each cohort are shown in Figure 2. The diversity of patient data is mainly reflected in various viral load levels to agree with prior studies that different viral load is associated with the severity of diseases or different factors such as age or sex of the patients [19].

**Figure 2:**
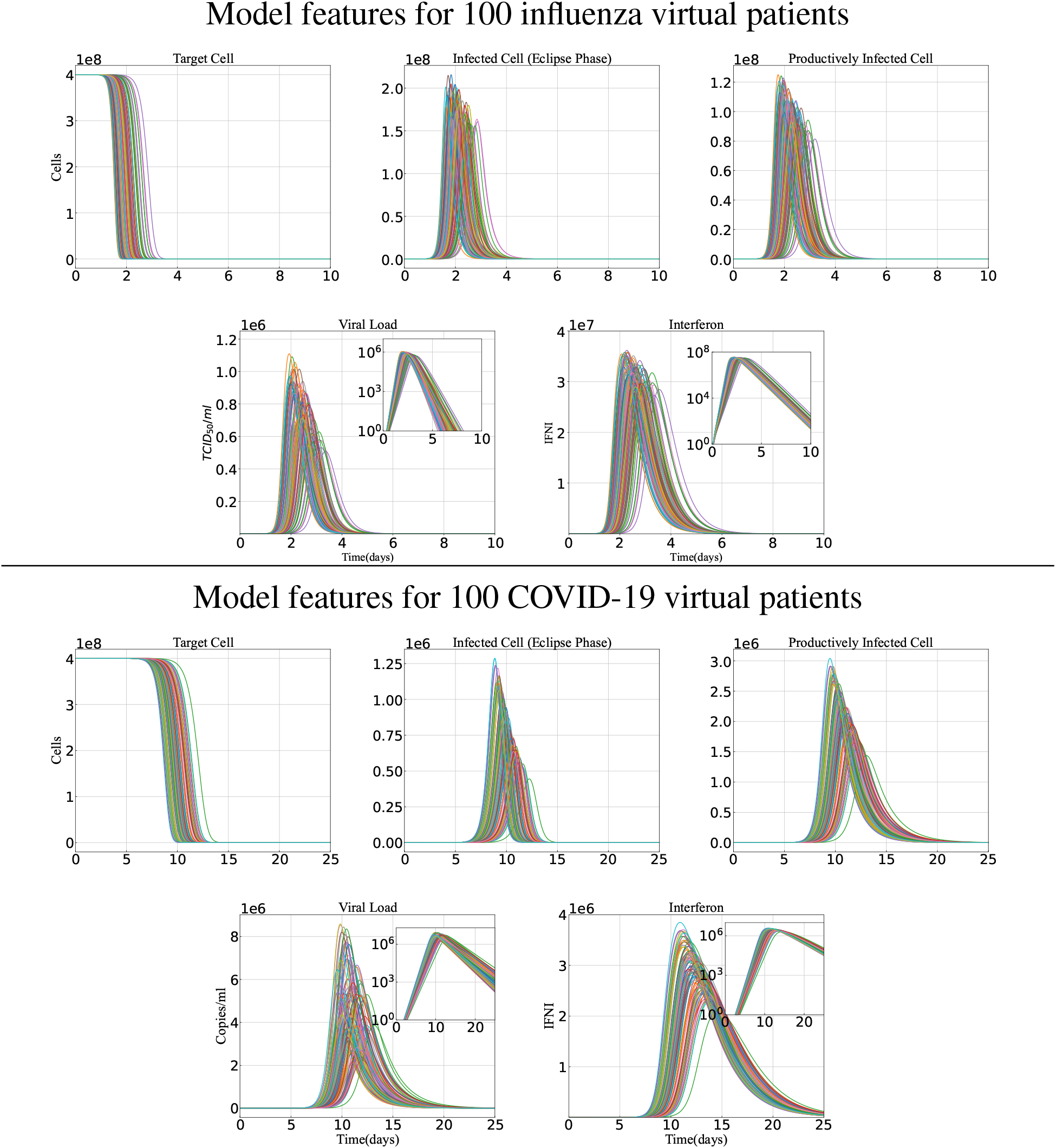
Cohort Dynamics. One hundred virtual patients are generated with different features of Target cells, infected/productively infected cells, viral load, and the only immune factor type I interferon for Influenza (upper two rows) and COVID-19 (lower two rows). Each solid curve with a different color represents a patient. The insets are in log scale.

#### Consistency of the data

Generating data with time consistency for different cohorts of infections is of great importance. Data inconsistency can lead to loss of information or biased results. Since the influenza mechanistic model predicts faster clearance of influenza-infected cells than SARS-CoV-2 [11], the infection period for influenza and COVID-19 patient dynamics are not the same, see Figure 2. Therefore we limited the consistency of flu/COVID-19 cohorts to have the same number of data points during the infection time. Hereupon, as an example, we divided the main infection period (i.e., [1 − 6] days for influenza patients and [10− 20] days for COVID-19 patients) into ten different sub-intervals with half-day length time steps for influenza patients and one and half-day length time steps for COVID-19 patients (see Figure 3). Hence, despite having different infection periods and time steps with different lengths to report the new virtual data point, the total number of data for the two different cohorts was the same.

**Figure 3:**
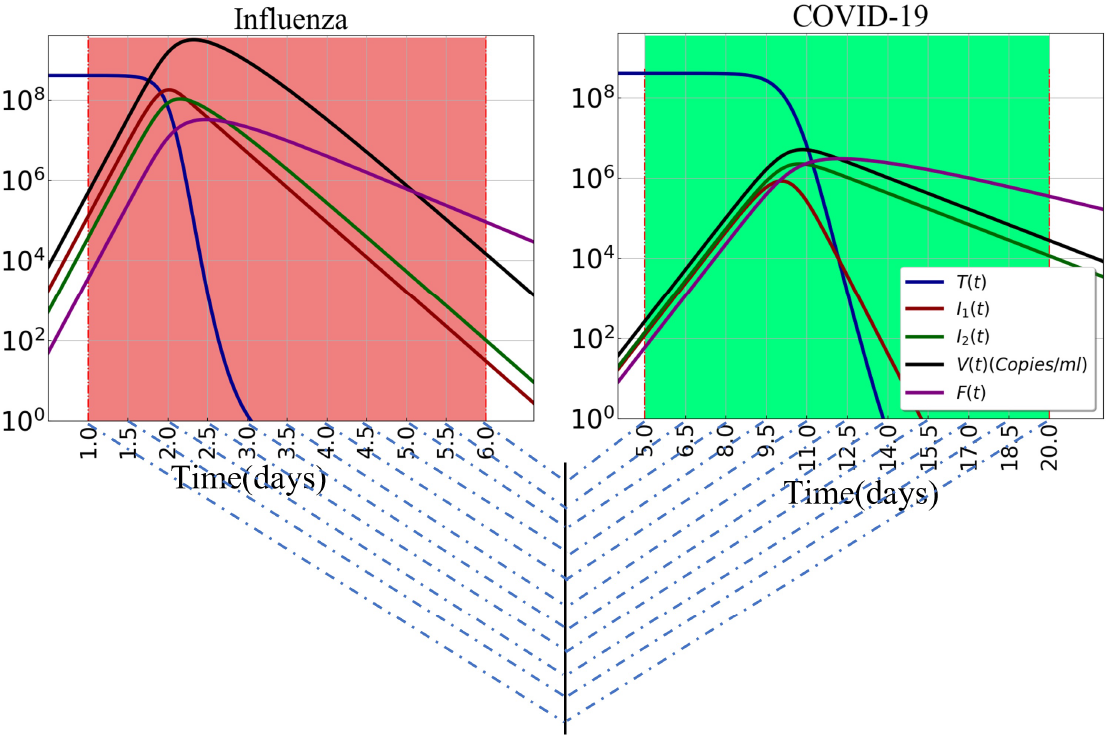
Consistency of the number of virtual data points during the time of infection. Dashed cross blue lines show eleven-time points of an influenza or COVID-19 patient.

In addition to the total infection period, we were also interested in studying the viral load dynamics in the early period of infection. The median incubation period for influenza A(B) virus is estimated to be 1.4(0.6) days, and for SARS-CoV-2 is around 5− 6 days [20]. Therefore, we assumed the time interval [0.9, 1.3] days for influenza, and [5 − 6.5] days for COVID-19 cohorts, corresponding to [10^2^− 10^4^]*Copies/ml* viral load. Dividing each interval into three different sub-intervals to get the time steps with length one-sixth of a day for Influenza and half a day for COVID-19 patients, we had four consistent data points for each patient.

### 2.3 Predictive Model Development

To distinguish between patients who encounter COVID-19 from those who are exposed to influenza, we developed a predictive model based on some biological feature selections. Accordingly, we adopted Logistic regression with *𝓁*_1_-regularization, referred to Lasso (stands for least absolute shrinkage and selection operator) Regression, as an appropriate technical classification. Lasso regression is widely used for many supervised classification problems based on the concept of probability [21]. It can simplify the model complexity by removing irrelevant features of the data set. Recently, this algorithm was used by Han et al. to find some additional novel immune features that accurately identified patients before the clinical diagnosis of preeclampsia [22].

Logistic regression, which is a special case of linear regression and used for binary classification, is defined by the following sigmoid function

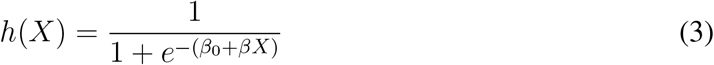

in which *X* is the (*n*×*p*) model feature matrix of *n* = 100 patients and *p* = 5 biological hallmarks. Defining the cost/objective (*C*) function of logistic regression in mean squared error format leads to a non-convexity that makes it difficult to optimally converge. Therefore, it is represented by the following equations

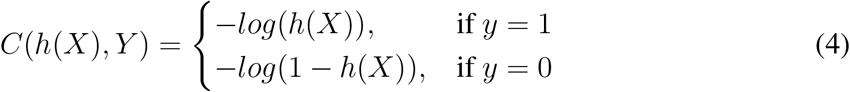

where *Y* is a binary response vector of outcome (CVOID-19 vs flu). Compressing the above two equations inside a single function, we have

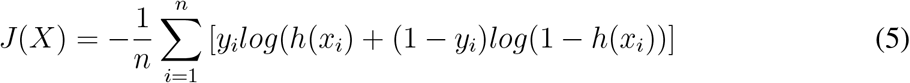

Replacing the sigmoid function from equation (3) and applying a penalty term equal to the absolute value of the magnitude of coefficients, we can reach the following objective function (after doing some mathematical simplifications) [22]

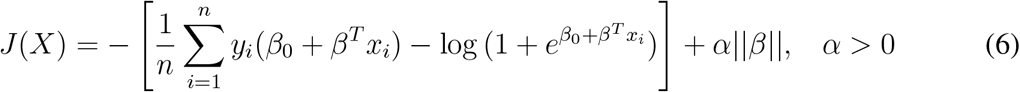

The penalty term which is called the *𝓁*_1_-regularization term is added to prevent data over-fitting. The model objective is to find a specific solution with a best-optimized cost function.

For model training and testing, we developed a *𝒦*-fold cross-validation strategy, which is a re-sampling method to evaluate machine learning models on a limited data sample. The procedure has a single parameter called *𝒦* which displays the number of groups that a given data sample is to be split into. As such, the procedure is often called *𝒦*-fold cross-validation. Therefore, our regression model is not tailored to a particular data set and is exposed to all available samples of a given subject in the training set. This approach implies that the training procedure was entirely blinded to the synthetic patient data sets, and ensures the presumed independence from any intra-subject correlations that are required for Lasso classification. We fixed the number of folds of the data as *𝒦*= 5. Running the analysis on each fold, the predicted outcome will be the one with the least estimated prediction error. The regularization parameter *α* is estimated by a cross-validation procedure.

#### 2.3.1 Evaluating Model Performance

The discriminating ability of the developed model in predicting patients with influenza from COVID-19 was evaluated using AUC (Area Under The Curve) ROC (Receiver Operating Characteristics) curve analysis. AUC - ROC curve is one of the most important evaluation metrics to visualize the performance of multi-class classification problems. ROC represents a probability curve of sensitivity 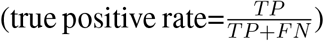 against 1-specificity 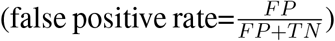 and AUC is a performance measure of discrimination. In the other words, the AUC score is a criterion that explains how well the model is capable of discerning different cohorts. Generally, an AUC closer to 1 indicates a better overall diagnostic performance of influenza classes as influenza or COVID-19 to COVID-19.

### 2.4 Model Interpretability

From [23, 24], “Interpretability” is the degree to which a human can understand the cause of a decision and consistently predict the model’s result. The higher the interpretability of a machine learning model, the better understanding of why certain predictions have been made. Interpretable machine learning models are beneficial to extract the relevant knowledge from relationships either contained in data or learned by the model [25, 26].

Here, we looked at the regularization path which is a plot of all coefficients values against the values of *α* in-*𝓁*_1_ penalization term, to see the behavior of the Lasso regression and interpret the prediction outcomes. The main purpose of Lasso regression is to classify groups of data by providing feature coefficients that can select the important features and maintain model regularization to avoid over-fitting the data. Therefore, the Lasso path can give us an idea of the feature’s importance.

## 3 Results

### 3.1 Prediction of Influenza versus COVID-19 infection

In this study we developed a classifier in the Lasso framework to identify patients with either influenza or COVID-19, based on four major entities of viral dynamics, {*T* (*t*), *I*_1_(*t*), *I*_2_(*t*), *V* (*t*) }, and one main factor of host immune response, type I interferon (*F* (*t*)), as the entry data features. The model was trained on data from one hundred virtual patient-level data in each infection cohort without noise, and it was externally validated on testing set with demographic noise (reflected in diverse viral load levels). Results in Figures 4, 5 and 6 reflect the Lasso predictions using the entire infection period (see Section 2.2). In Figures 4 and 5, two-dimensional scatter plots are used to compare ground truth to regression predicted values based on all model features. The hue spectrum from light to dark illustrates the probability of being in the influenza (blue) or COVID-19 (red) group. In the other words, the darker the colors, the better the prediction. Considering three attributes in the data, the predicted outcomes are improved. This is shown in three-dimensional scatter plots in Figure 6 of the ground truth and regression predicted values. ROC AUC=95% indicates a satisfactory performance of the model to distinguish between COVID-19 and influenza patients, see figure 7 for more details.

**Figure 4:**
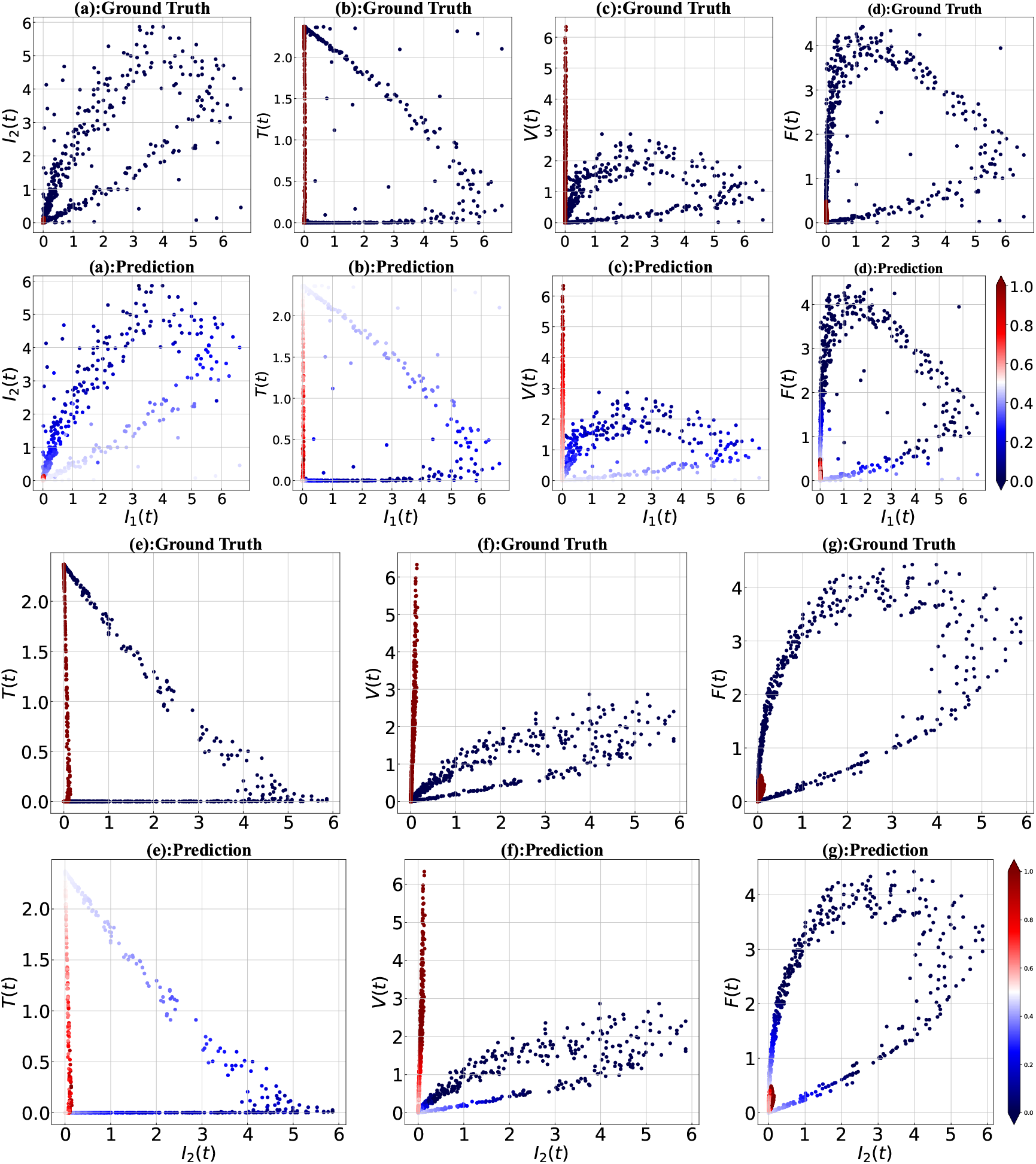
Two-dimensional scatter plots of ground truth and regression predicted values based on model features. Classification of the data was done for: *I*_2_ versus *I*_1_ in panels (a), *T* vs. *I*_1_ in panels (b), *V* vs. *I*_1_ in panels (c), *F* vs. *I*_1_ in panels (d), *T* vs. *I*_2_ in panels (e), *V* versus *I*_2_ in panels (f), and *F* versus *I*_2_ in panel (g). Color denotes the patient probability of being in the influenza (blue color scheme) or COVID-19 (red color scheme) cohorts. Data points, corresponding to each model feature, are rescaled by dividing by their standard deviations.

**Figure 5:**
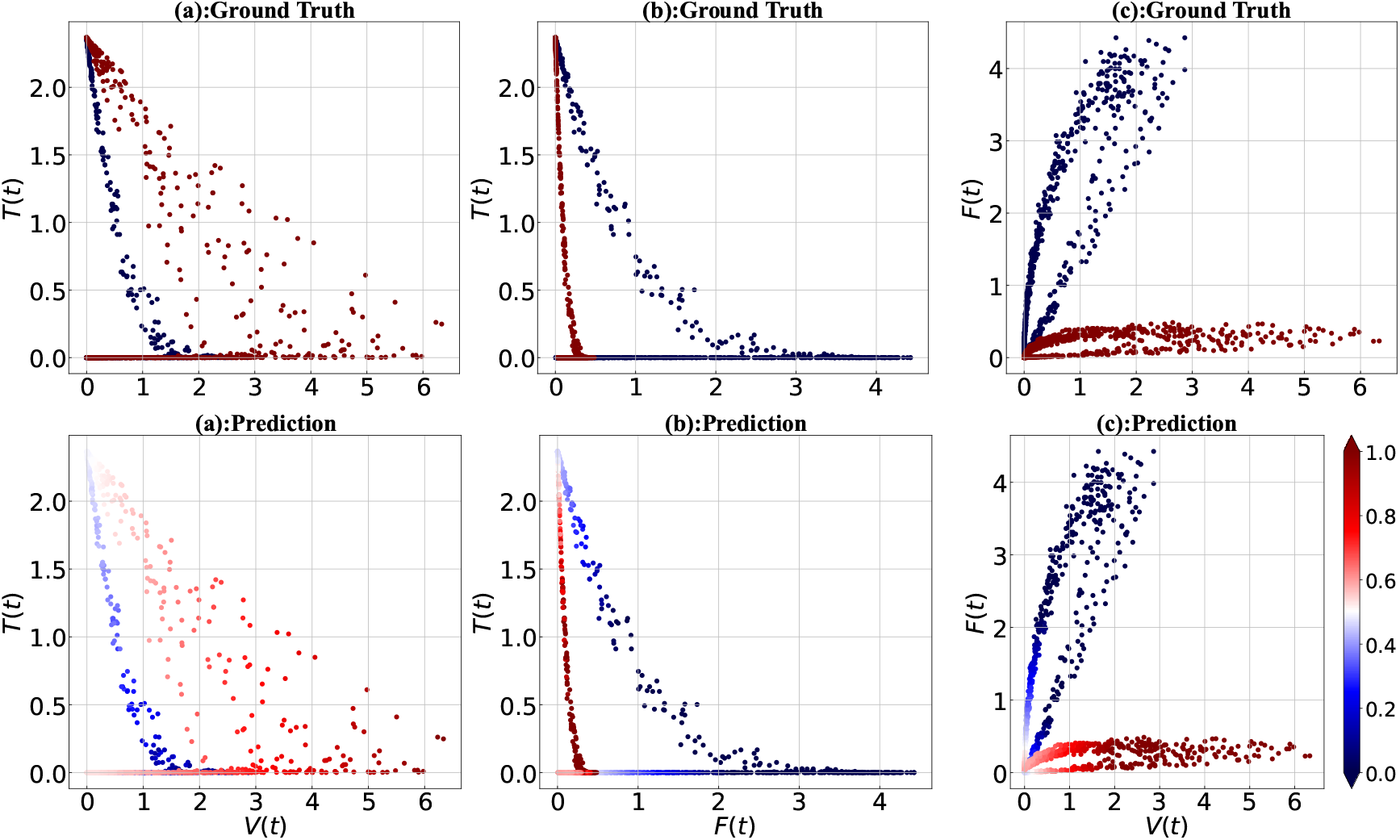
Two-dimensional scatter plots of the ground truth and regression predicted values for three model features *T, V, F*. Classification of the data was done based on: *T* versus *V* in panels (a), *T* versus *F* in panels (b), and *F* versus *V* in panels (c). Color denotes the patient probability of being in the influenza (blue color scheme) or COVID-19 (Red color scheme) cohorts.

**Figure 6:**
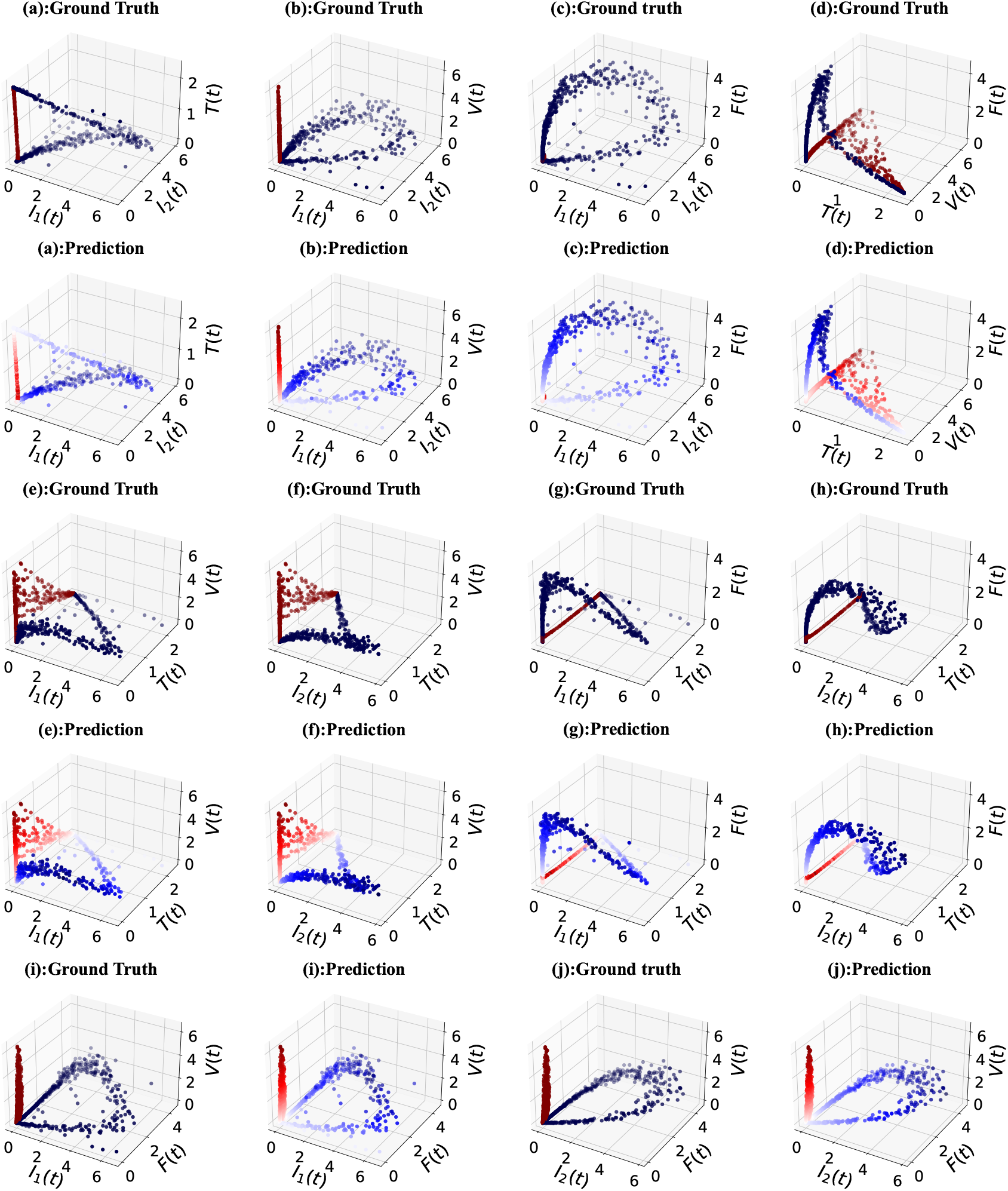
Three-dimensional scatter plots of the ground truth and regression predicted values based on all model features. Classification is based on *I*_1_, *I*_2_, *T* in panels (a), *I*_1_, *I*_2_, *V* in panels (b), *I*_1_, *I*_2_, *F* in panels (c), *T, V, F* in panels (d), *I*_1_, *T, V* in panels (e), *I*_2_, *T, V* in panels (f), *I*_1_, *T, F* in panels (g), *I*_2_, *T, F* in panels (h), *I*_1_, *F, V* in panels (i), and *I*_2_, *F, V* in panels (j). Shades of blue (red) indicate influenza (COVID-19) group patients. Data points are dimensionless by dividing by the corresponding standard deviations.

**Figure 7:**
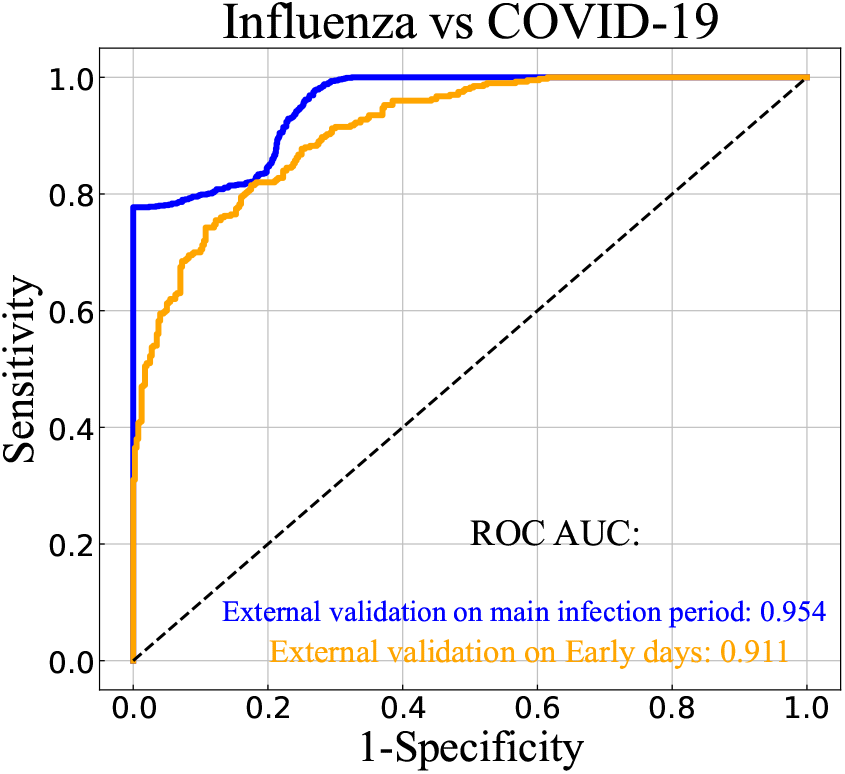
Receiver Operating Characteristic curve (ROC) of influenza vs COVID-19 patients. Area under the ROC curve indicates the predictive performance of the model between COVID-19 and influenza encounters on the external validation test during the main infection (blue curve) and early days (orange curve). The black dashed line in the diagonal has a ROC AUC of 0.5.

#### Early days of infection

We examined the model prediction for the data generated at the early days of infection after the incubation period. The results are shown in Figure 8 based on the model features. From the figure, we can see that there are some mispredictions, for small values of *I*_1_(*t*), *I*_2_(*t*), *V* (*t*), and *F* (*t*), especially when *I*_2_(*t*) is plotted as a function of *I*_1_(*t*) or *V* (*t*) is plotted in terms of *I*_2_(*t*). In the other words, for this range of values, the influenza patients were misdiagnosed with COVID-19. In an attempt to find the reason, we compared correlations between the different variables in our model – see Figure 9). Here, we see small regions of overlap between influenza and COVID-19 models. Accordingly, the compatibility of the results between the two infections may lead to some overlaps in the model predictions. However, the ability of the model in the prediction of infections when the patients were monitored by *V* (*t*)/*F* (*t*) as a function of *I*_1_(*t*), panels (b) and (c), or *F* (*t*) in terms of *I*_2_(*t*)/*V* (*t*), panels (e) and (f), can be satisfactory, and thus can serve as benchmarks for clinical diagnosis. The model had a ROC AUC of 91% on the external validation data set for early infection – see Figure 7.

**Figure 8:**
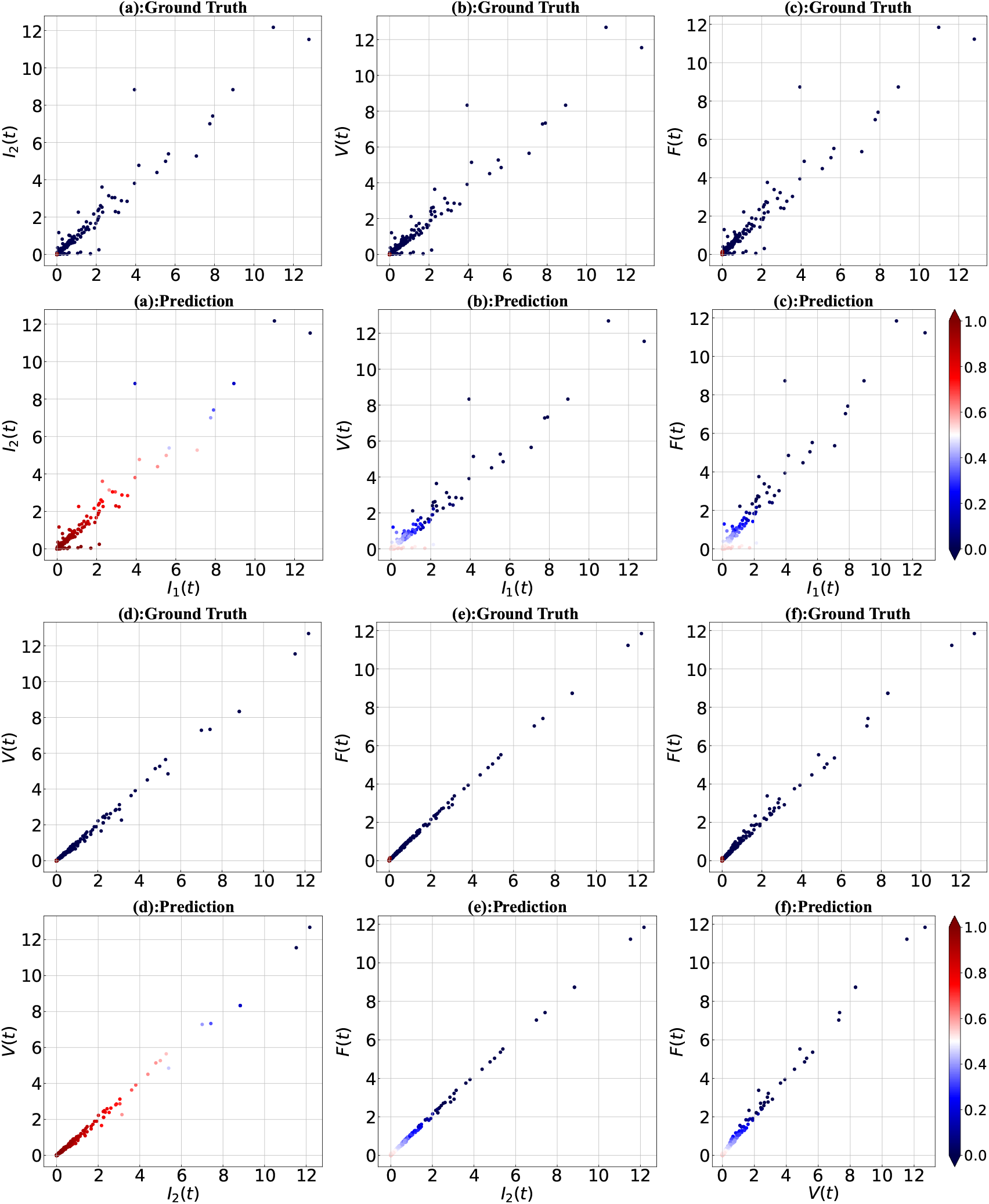
Early days of infection. Two-dimensional scatter plots of the ground truth and regression predicted values based on model features are shown. Classification is based on *I*_1_, *I*_2_ in panels (a), *I*_1_, *V* in panels (b), *I*_1_, *F* in panels (c), *I*_2_, *V* in panels (d), *I*_2_, *F* in panels (e), and *V, F* in panels (f). Shades of blue (red) indicate influenza (COVID-19) group patients. Data points are dimensionless by dividing by the corresponding standard deviations.

**Figure 9:**
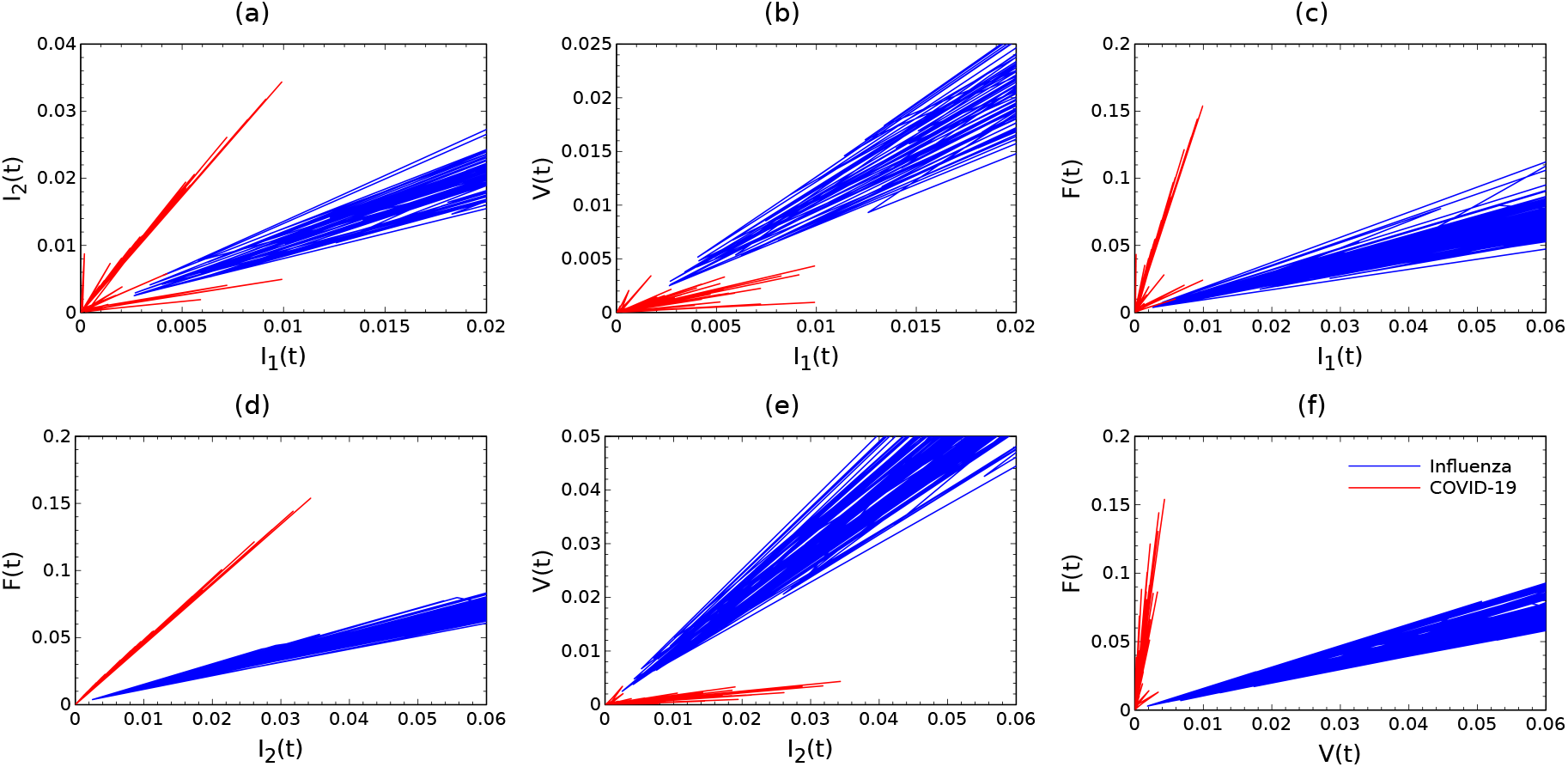
Comparison of in-host measurements, *{T, I*_1_, *I*_2_, *V, F}*, between influenza and COVID-19 virtual patients where plotted as a function of each other. Blue(red) solid lines represent the ratio of the features for one hundred influenza (COVID-19) patients. Data points are divided by the corresponding standard deviations for each feature.

### 3.2 Significance of the features

To investigate the importance of various data features we created our *𝓁*_1_-regularization path, which was the best way to see the behavior of the Lasso regression. The regularization path is a plot of all coefficient values in terms of the regularization parameter. Figure 10 illustrates the selection path of each feature with its corresponding coefficient in terms of the logarithm of the regularization parameter *α*. For each value of *α*, the path method on the Lasso object returns the coefficients that solve the logistic regression problem with that parameter value. The optimal value of − log(*α*) was estimated at around 3.25 for the test set distributed over the entire infection course, and 3.04 when the early days of infection were studied. The results suggested a higher coefficient value for viral load *V* (*t*) and productively infected cells *I*_2_(*t*) compared to the other features.

**Figure 10:**
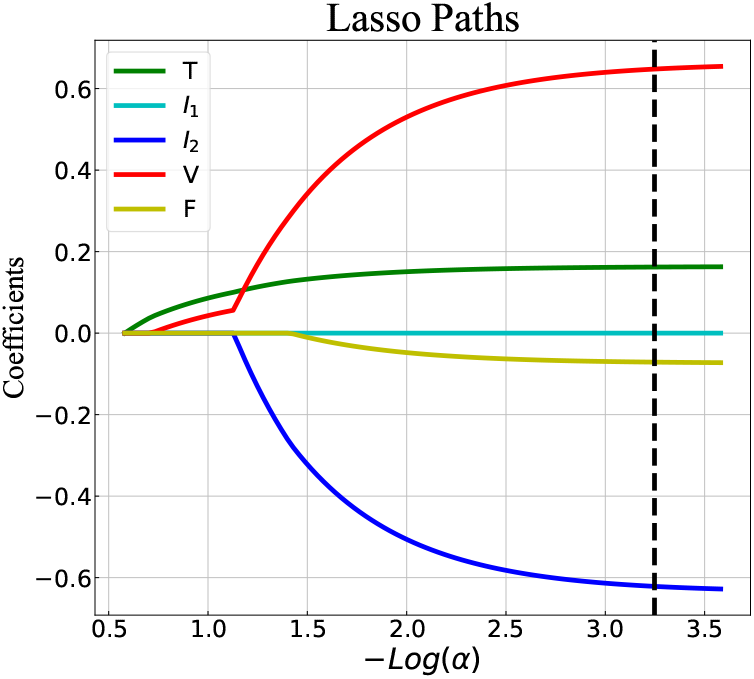
Lasso coefficients of five sample features, *{T, I*_1_, *I*_2_, *V, F}*, as a function of the logarithm of regularization parameter, − log *α*. Each colored line represents the value taken by a different coefficient in the optimization objective for Lasso. The black dashed line indicates the selected regularization parameter with the value of − log(*α*) ∼ 3.25. This number was ∼ 3.04 with the same Lasso Paths when the early days of the infection period were considered.

## 4 Discussion

This study presents a machine learning model to effectively classify influenza and COVID-19 virtual patients using in-host patient data. Our model employed a Lasso regression classifier trained to identify between two hundred patients, highlighted by a ROC AUC of 95%. Using the existing within-host models, We generated synthetic data with five in-host measurements including target cells, eclipse phase, and productively infected cells, viral load, and type I IFN. Analyzing the feature importance revealed that the viral load and the productively infected cells are the most important components to determine if a patient is infected by influenza or SARS-CoV-2.

While our machine learning model was built on the synthetic data distributed during the main infection period, it ascertained a good performance (ROC AUC = 91%) even for the early days, once after the incubation period. However, there are some exceptions for the small values of in-host features where the influenza patients are misdiagnosed by COVID-19 for the early days of infection. The reason was explained by the fact that during the early days of the infection, influenza and COVID-19 patients have comparable in-host measurements that lead to some errors in discriminating the patients. This is interpreted as a limitation of our model and can be a future extension of developing dynamic models which take more immune entities into account and end in a better classifier.

Our model was trained and successfully evaluated on synthetic data. The model, however, could be applied to animal or human clinical data. This could be useful, for example, if a clinical trial is complicated by the existence of an infectious disease with similar infection characteristics. The model could be applied as a low-cost classification system that would not require expensive virus typing procedures and could rely solely on viral load and interferon measurements. We note that studies like [9] that focus analysis on demographic and observational data can be cheaper to conduct, but these data can also be subject to inconsistencies and bias, affecting classification outcomes. In a future study, we will expand our analysis to a model of in-host measurements and observational data to determine if specific combinations of in-host and observational data that best classify influenza and COVID-19 infections differ.

Our machine learning model was developed in the Lasso framework. Ridge regression could also be employed, and require only small changes to our method to include this. We find that the model demonstrated a satisfactory performance by using a Ridge regression classifier – (ROC AUC= 95%) for the main infection period, and (ROC AUC= 89%) for the early days of infection.

## Data Availability

All data produced in the present study are available upon reasonable request to the authors

